# Trends and Adverse Outcomes of Xylazine Misuse: A Digital Surveillance Study

**DOI:** 10.1101/2024.01.31.24302075

**Authors:** Akshaya Srikanth Bhagavathula, Wafa A. Aldhaleei, Seongjin Kim

## Abstract

**Background:** Xylazine is an animal tranquilizer without approved medical use in humans that is increasingly being misused as an adulterant in illicit drugs. This study aimed to characterize national trends and adverse outcomes associated with the emerging misuse of xylazine using digital surveillance data.

**Methods:** We examined online search trends and social media discussions related to xylazine misuse in the U.S. from 2019-2023 using Google Searches data and conducted Joinpoint regression to assess trends. We also examined social media attention using Almetric attention score and analyzed reports on xylazine in the FDA Adverse Event Reporting System (FAERS) through September 2023 using logistic regression.

**Results:** Our analysis revealed an overall increasing trend in online searches for “xylazine” nationally, with an average monthly percentage change of 4.6% (95% CI: 3.9–5.1, *P*_trend_ <0.001), indicating growing awareness. On social media, mentions of xylazine rose exponentially starting in late 2021. Analysis of the FAERS data identified 94 reports of adverse events related to xylazine, most of which involved men (70.2%), with a mean age of 36.5 ± 14.3 years. Alarmingly, these xylazine-linked adverse events had an 87.2% fatality rate, which increased over 40-fold with concurrent fentanyl use (reported OR: 40.5, 95% CI: 4.0–407.4, *P*=0.002).

**Conclusions:** These findings underscore the urgent need for greater public health awareness, harm reduction strategies, and enhanced surveillance targeting the worsening xylazine addiction and overdose crisis.

## Introduction

The ongoing opioid epidemic continues to exact a devastating toll in the United States, evidenced by surging opioid overdoses and rising mortality [1]. Even as public health measures ramp up to curb this crisis, the rapid emergence of novel psychoactive substances further compounds risks among people who use drugs. In particular, the veterinary tranquilizer xylazine has infiltrated illicit drug supplies as a new adulterant, posing alarming, unanticipated threats to an already vulnerable population [2].

Originally approved as an analgesic, sedative, and muscle relaxant for animal use, xylazine is increasingly being misused as an emerging new psychoactive substance [3]. Human consumption of xylazine was first described in Puerto Rico during the early 2000s, often in combination with opioid use, though it has spread stateside over the past decade [4-6]. Case reports and local surveillance data indicate that xylazine is increasingly being used as an adulterant in street supplies of opioids like heroin or fentanyl, heightening overdose risks [7-10].

Xylazine has central nervous system (CNS) depressant effects and may potentiate and prolong opioids effects, instead compounding respiratory depression and fatal overdoses when mixed with other sedatives [11]. While naloxone can reverse opioid overdoses, it has no impact on the effects of xylazine, which cannot be readily reversed or treated [12]. Consequently, xylazine is exacerbating overdoses associated with injection drug use across multiple states, though national surveillance data characterizing the scope and public health impacts have been notably absent.

Quantifying the rising crisis associated with improper xylazine misuse is critically needed to guide public health strategies targeting prevention and harm reduction. Offering timely insights into a rising threat facing people who use drugs, providing an empirical foundation to guide public health measures addressing the complex challenges at the nexus of the evolving opioid and novel psychoactive substance crises.

Online searches and social media discussions are increasingly used for public health surveillance to identify emerging issues and monitor trends over time [13,14]. However, little attention has focused specifically on modeling opioid-related internet search patterns as a barometer of evolving misuse threats [15-17]. With recent U.S. Drug Enforcement Administration warnings highlighting sharp increases in fentanyl adulterated with the veterinary tranquilizer xylazine [18], digital surveillance represents an invaluable tool to track this worsening risk facing people who use drugs.

Given the lack of empirical evidence to better understand the scope of the xylazine abuse problem, examining online searches and social media discussions can serve as a proxy for interest and information-seeking behavior. Thus, this study leverages digital surveillance from online search trends and social media monitoring through Google Health Trends and Almetric analysis to empirically characterize the trajectory of the emerging xylazine adulterant crisis and associated health impacts in the U.S. In addition, we also analyzed the FDA Adverse Event Reporting System (FAERS) database to investigate the occurrence of adverse health events associated with the non-medical utilization of xylazine.

Quantifying the rising crisis associated with improper xylazine misuse is critically needed to guide public health strategies targeting prevention and harm reduction. Offering timely insights into a rising threat facing people who use drugs, providing an empirical foundation to guide public health measures addressing the complex challenges at the nexus of the evolving opioid and novel psychoactive substance crises.

## Methods

### Study design and data sources

This was a cross-sectional infodemiology study conducted using three major data sources: 1) Google online searches, 2) Almetrics database explorer, and 3) the FAERS database.

#### 1. Google online searches

Google Trends and wordstream webpages were used to identify the top search key terms and queries associated with ‘xylazine’ and their respective search volumes from 2019 to 2023. The Google Health Trends API (GHT-API), which returns a 10-15% sample of raw search data as probability values per million searches, was then leveraged to obtain normalized search probabilities for these terms at both the national and state levels. To minimize missing data, a resampling methodology adapted from prior research involved extracting 30 unique repeated samples over the study period for key search terms including *xylazine, tranq, rompun, zombie drug*, and *traq dope*. This approach provided robust estimates of search trends associated with emerging xylazine misuse over the study period, mitigating the influence of idiosyncratic single-date extractions. Further details on the GHT-API methodology have been described previously [19,20].

#### 2. Social media attention

The Altmetric attention score is an automatic algorithm created by Altmetric that calculates weighted scores reflecting the social media attention received by published articles [21]. Altmetric began tracking online attention for published research in 2011 across Twitter, Facebook, policy documents, news media, blogs, Mendeley, Reddit, Wikipedia, Google+ and other sources [22]. For published articles mentioning “xylazine”, additional variables collected included the Altmetric score, number of tweets, Facebook mentions, and news stories referencing the article. Mentions were procured from the Altmetric attention score reported on journal websites, which use the same consistent Altmetric algorithm. This approach allowed quantitative characterization of social media and news attention towards emerging literature related to xylazine misuse over the study timeframe.

#### 3. FAERS database

The U.S. Food and Drug Administration FAERS is a national database comprising adverse drug event and medication error reports submitted to the FDA through the MedWatch reporting program [23]. FAERS represents one of the largest U.S. government databases monitoring post-marketing drug safety. All adverse events related to xylazine exposure among U.S. cases from inception through September 30, 2023 were downloaded, cleaned and harmonized. This FDA FAERS data enabled examination of the demographics, clinical effects, and outcomes associated with emerging xylazine misuse at a national level based on empirical adverse event reports over the study period.

### Statistical analysis

A range of descriptive and inferential statistical techniques used for analyses. Online search trends were visually depicted using plots of monthly search volumes during the study period across U.S. states using the R package geofacet version 0.2.0 [24]. Social media mentions related to xylazine across platforms including tweets, news stories, Facebook posts, Reddit discussions and other sources were aggregated and presented visually using a line graph depicting temporal trends in total mentions over the study period. Joinpoint regression analysis was performed using Joinpoint software version 5.0 to identify significant changes over time in search trends [25]. This approach pinpoints trend inflection points (“joinpoints”) and estimates the regression function between joinpoints [26]. The monthly percentage change (MPC) summarized linear trends in search volumes. Models incorporated up to 3 joinpoints and were selected based on the best model fit. Joinpoint regression was applied to Google searches for “xylazine” in the U.S. from 2019-2023 to quantify monthly percentage changes with 95% confidence intervals (CIs). The average MPC yielded the overall percentage increase in searches over the study period. For the FDA FAERS data, xylazine-related adverse event cases were summarized using descriptive statistics including means/SD for continuous variables and frequencies/percentages for categorical outcomes. Multivariable logistic regression analysis was conducted using Stata v15.1 (StataCorp) to estimate adjusted reported odds ratios (ROR) with 95% CIs for mortality associated with different xylazine exposure conditions, controlling for potential confounders. A *P*-value ≤ 0.05 was considered statistically significant.

## Results

Examination of Google search trends for “xylazine” revealed an overall increasing interest in this emerging drug of misuse both nationally and at the state level from 2019 to 2023. Nationally, the average monthly percentage change (AMPC) in searches was 4.6% (95% CI: 3.9-5.1, *P*_trend_ <0.001), indicating a significant rising trend over time. Segmented regression identified an inflection point in December 2022 when searches began accelerating more rapidly through March 2023 (MPC 101.2%, 95% CI: 57.3-121.5, *P*_trend_ <0.001).

A significant increasing trend in online searches for xylazine were observed over the study period across all U.S. states. The most dramatic increase began in January 2022 and continued rising through September 2023 in every state, indicating growing awareness and concern. More details provided in **Figure 1**.

**Figure 1:**
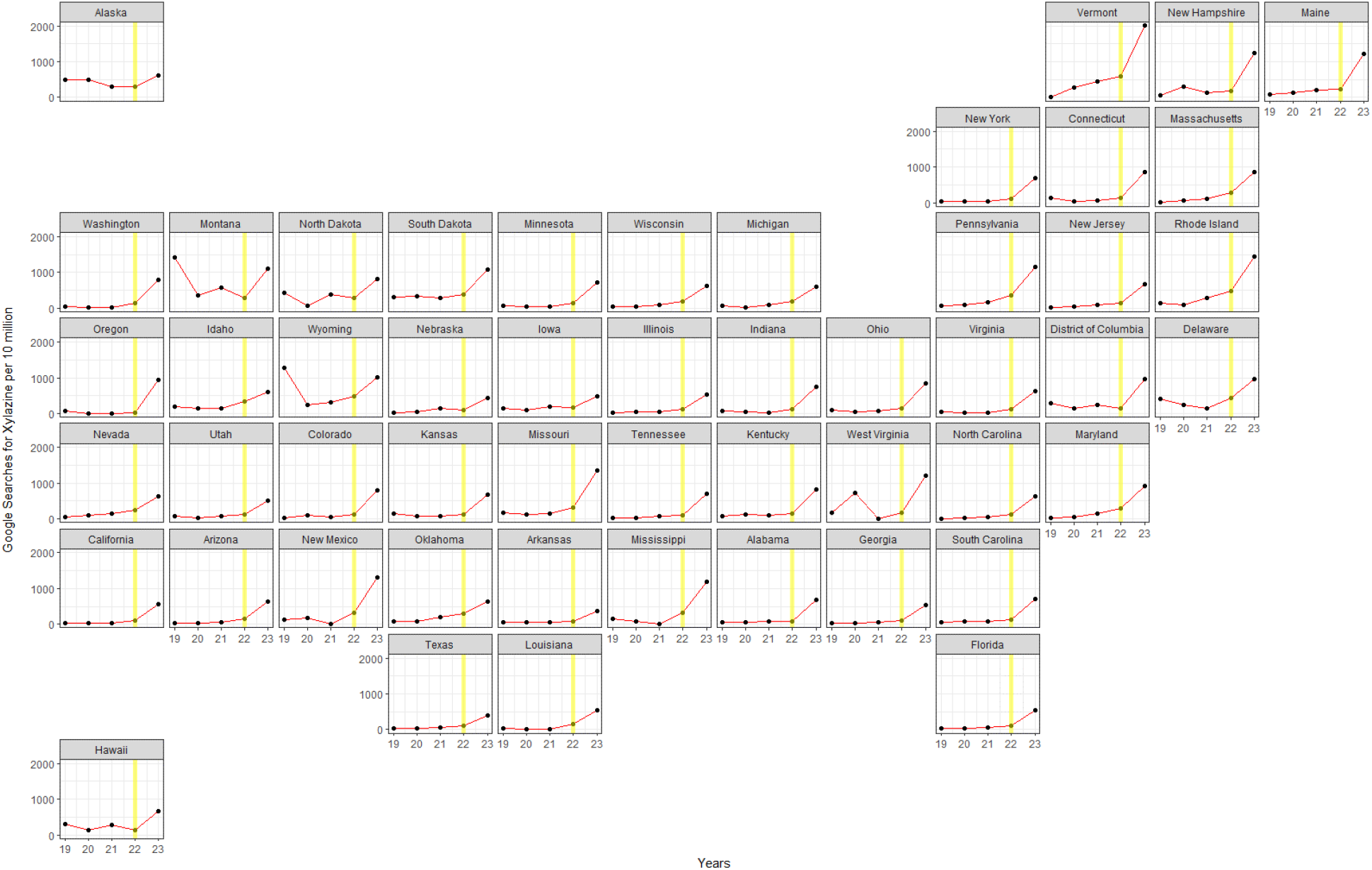
Internet Searches on Xylazine in the U.S. from January 1, 2019 – September 30, 2023. Monthly searches (red line), years (red dots) and yellow line (starting from January 2022).

Examination of social media mentions and discussions pertaining to xylazine showed relatively flat levels from 2020 through most of 2021. Starting in late 2021, there was growing interest and discussion of xylazine, with exponential increases on Twitter and in news media. Mentions peaked in mid-2022 as public attention to this issue escalated (**Figure 2**).

**Figure 2:**
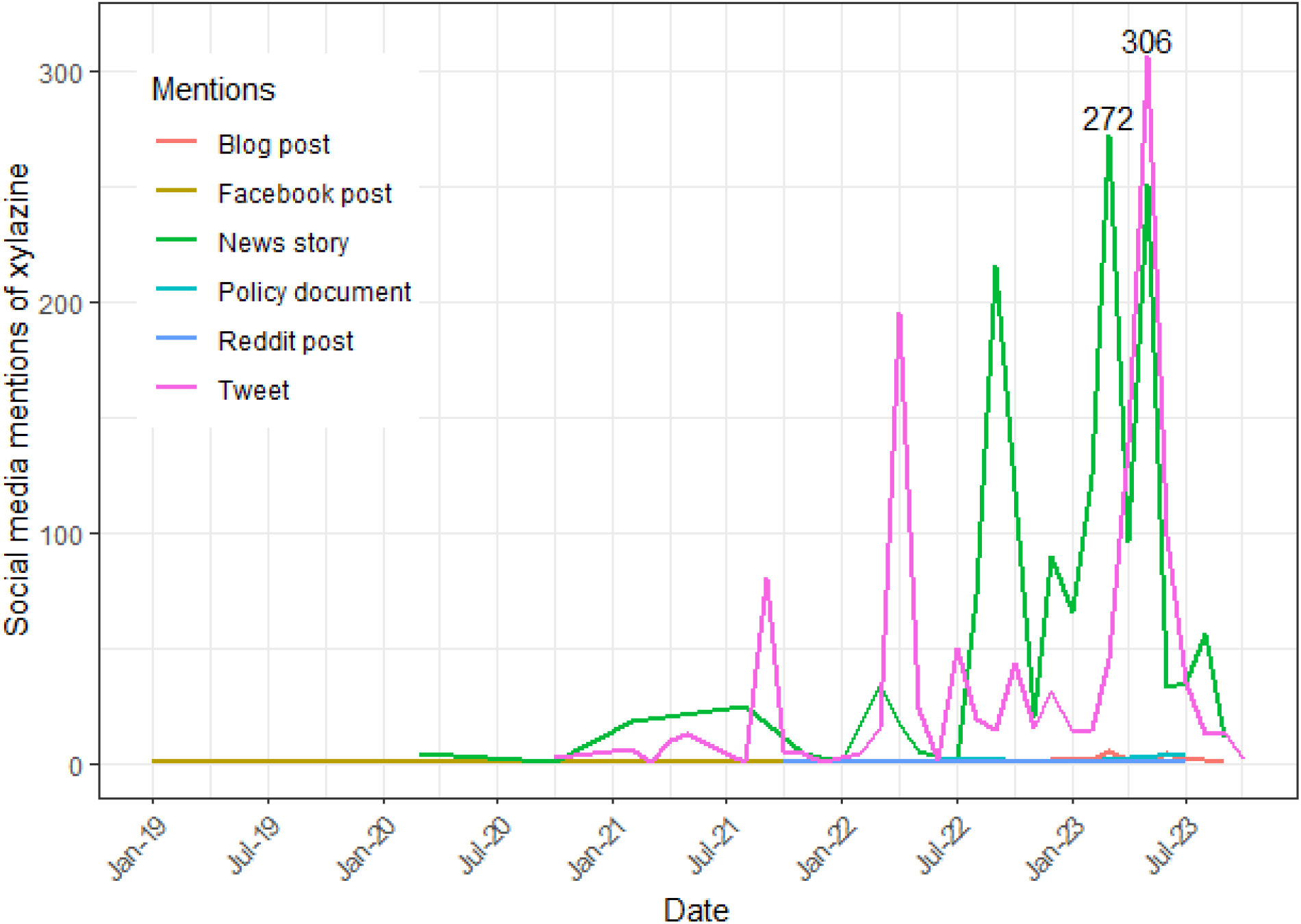
Mention of xylazine in various social media posts from 2020 – 2023 (September)

Our analysis of the FAERS database identified 94 reports of adverse events related to xylazine, providing demographic details and outcomes for affected individuals. The majority of adverse event reports involved men (70.2%) with a mean age of 36.5 ± 14.3 years (see Table 1). Alarmingly, these xylazine-linked adverse events had an 87.2% fatality rate. Moreover, concurrent exposure with the synthetic opioid fentanyl further increased the odds of xylazine-related mortality by over 40-fold compared to xylazine alone (adjusted ROR: 40.5, 95% CI: 4.0 – 407.4; *P* = 0.002).

**Table 1:**
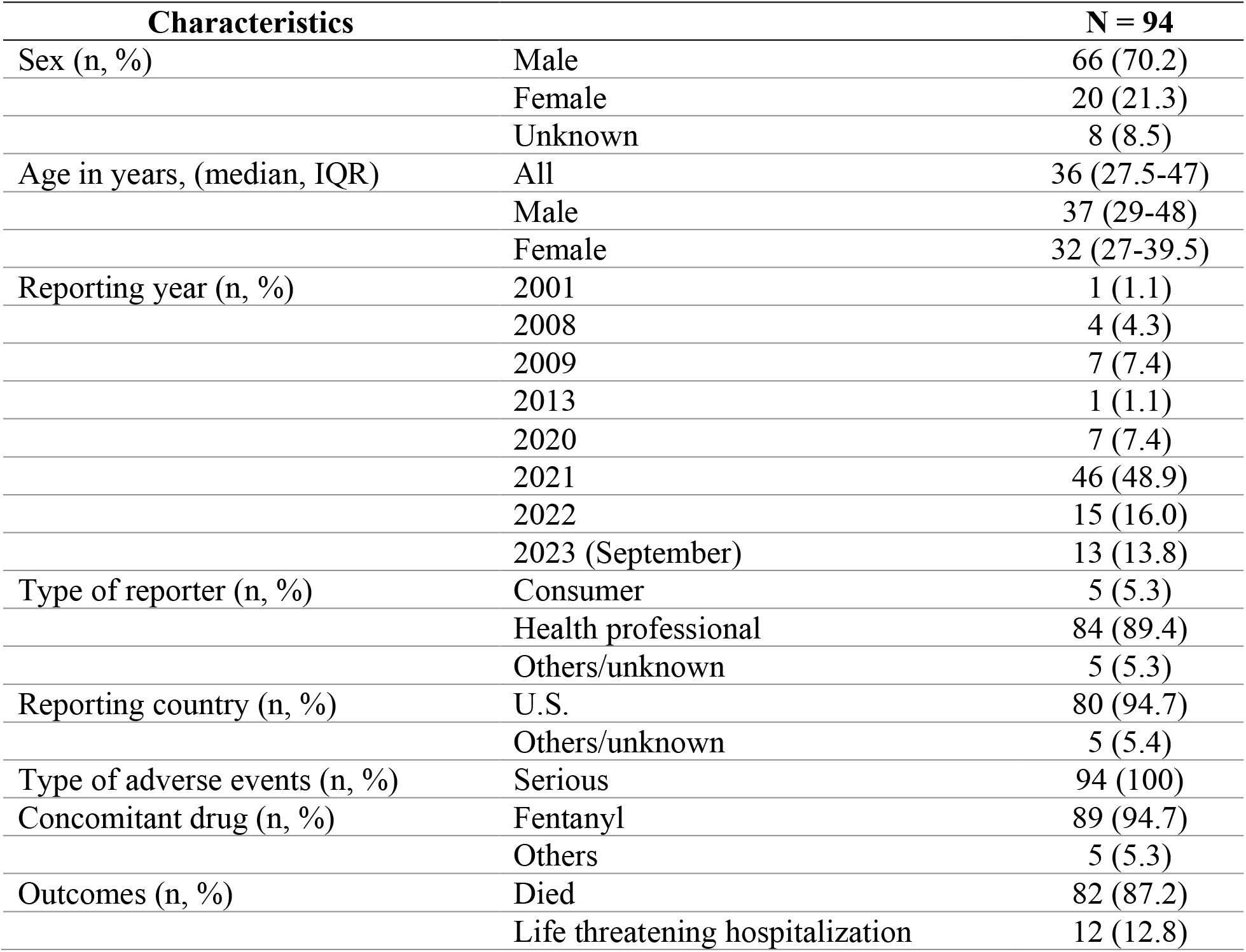
Characteristics of adverse events reports associated with xylazine from 2001 to 2023.

## Discussion

Our study provides a comprehensive analysis of the rising misuse of xylazine, an animal tranquilizer, with illicit drugs, using digital surveillance data. The findings reveal an alarming increase in xylazine misuse and its associated harms, particularly when used concurrently with synthetic opioids like fentanyl. The analysis of the FAERS database identified 94 reports of adverse events related to xylazine, with a majority involving men (70.2%) and a mean age of 36.5 years. The fatality rate associated with these xylazine-linked adverse events was a staggering 87.2%. Furthermore, concurrent exposure with fentanyl increased the odds of xylazine-related mortality by over 40-fold compared to xylazine alone. These findings align with the existing literature on the misuse of xylazine and its associated harms [12,27,28]. However, our study adds to this body of knowledge by providing a detailed analysis of national trends and adverse outcomes associated with xylazine misuse, using multiple data sources and advanced statistical methods.

Our findings possibly reflect growing public recognition, apprehension, and focus towards the emerging xylazine abuse crisis as it unfolded in real-time, offering clues even earlier than traditional public health data collection systems [12,27]. Notably, adverse events identified from FAERS showed the extreme lethality of xylazine misuse, particularly when adulterating opioids like fentanyl. While these adverse event reports likely underestimate the true mortality burden, they offer valuable insights into the demographics and risks of xylazine misuse. Our digital surveillance approaches efficiently synthesized real-time data on an emerging public health threat.

Our findings should be interpreted within the broader landscape of public health efforts and research targeting the syndemic of opioid use disorder, injection drug harms, and overdose mortality in the United States. As illicit supplies transform, often faster than surveillance systems, the rapid adoption of multi-modal digital tracing here presents the value of these methods for tracking emerging threats. However, work by Palamar and colleagues [28] highlights geographic and demographic blind spots that persist, affirming needs to expand surveillance through linkage with medical examiner data and wastewater analysis capturing localized community impacts. Enhanced coordination across public health, law enforcement, and health systems is crucial to deter diversion of veterinary tranquilizers while implementing evidence-based harm reduction strategies. As users increasingly face polysubstance exposures, establishing patient-centered approaches remains critical — understanding distinct clinical effects, risk profiles, and experiences to better prevent and treat substance use disorder in the face of a rapidly evolving crisis.

Our investigation emphasizes the urgent need for greater public health awareness, harm reduction strategies, and enhanced surveillance targeting the heightening xylazine abuse crisis, given the sharp rise in online interest and adverse outcomes associated with misuse of this veterinary tranquilizer, particularly when adulterating illicit fentanyl [12]. Key areas for future research include potential interventions to deter xylazine adulteration and mitigate harms, characterizing clinical effects and risk factors, geographic and demographic surveillance to identify hotspots, and examining public perceptions [28], and examining public perceptions to guide education and messaging. A multifaceted public health and research agenda is critically needed to curb the escalating mortality burden associated with improper xylazine use.

Our study has several strengths. To the best of our knowledge, this is the first study that examined infodemiology by combining Google online searches, Almetrics database explorer, and the FAERS database. Moreover, the Almetric tracking evaluation was done using several platforms and social media sites, providing a broad characterization of online discourse. Furthermore, the rigorous resampling methodology applied to Google searches helped minimize missing data. One limitation is the cross-sectional nature of the study hindering the ability to establish causality; however, this investigation provides valuable insights into the xylazine crisis for future studies. The FAERS adverse events reports may underestimate the mortality burden due to sampling bias and missing data; therefore, comprehensive ongoing surveillance methods are warranted to fully evaluate xylazine use-associated morbidity and mortality.

Future research should aim to understand why xylazine misuse is increasing and to develop effective interventions. For instance, qualitative studies could explore the reasons behind xylazine misuse from the perspectives of those using it, while intervention studies could rigorously test the effectiveness of different harm reduction strategies in at-risk communities. Additionally, continued surveillance is needed to track emerging geographic hotspots as illicit drug supplies evolve. Furthermore, monitoring unpopular groups on fringe social media platforms could provide valuable surveillance capabilities for tracking the online illegal marketing and sales of xylazine.

## Conclusions

In summary, our study provides compelling evidence of the rising misuse of xylazine and its associated harms. These findings highlight the imperative necessity for pronounced public health awareness and harm reduction strategies, and improved surveillance aiming to contain xylazine overdose crisis. Future studies are warranted to evaluate clinical effects and risk factors associated with xylazine use.

## Data Availability

All data produced in the present study are available upon reasonable request to the authors

## References

1. Spencer MR, Cisewski JA, Warner M, et al. Drug Overdose deaths involving Xylazine: United States, 2018–2021. Vital Stat Surveil Report 2023, Report 30. Available from: https://www.cdc.gov/nchs/data/vsrr/vsrr030.pdf.

2. Ruiz-Colón, K., Chavez-Arias, C., Díaz-Alcalá, J.E., et al. Xylazine intoxication in humans and its importance as an emerging adulterant in abused drugs: a comprehensive review of the literature. Forensic Sci Int. 2014;240: 1–8. 10.1016/j.forsciint.2014.03.015.

3. Friedman, J., Montero, F., Bourgois, P., et al. Xylazine spreads across the US: A growing component of the increasingly synthetic and polysubstance overdose crisis. Drug Alcohol Depend. 2022;233, 109380. 10.1016/j.drugalcdep.2022.109380.

4. Torruella RA. Xylazine (veterinary sedative) use in Puerto Rico. Subst Abus Treat Prev Policy. 2011;6 (1), 7. 10.1186/1747-597X-6-7.

5. Rodríguez N, Vargas Vidot J, Panelli J, et al. GC-MS confirmation of xylazine (Rompun), a veterinary sedative, in exchanged needles. Drug Alcohol Depend. 2008;96 (3),290–293. 10.1016/j.drugalcdep.2008.03.005.

6. Reyes JC, Negrón JL, Colón HM. The emerging of xylazine as a new drug of abuse and its health consequences among drug users in Puerto Rico. J. Urban Health. 2012;89 (3), 519–526. 10.1007/s11524-011-9662-6.

7. Mulders, P., van Duijnhoven, V., Schellekens, A. Xylazine Dependence and Detoxification: A Case Report. Psychosomatics. 2016;57(5), 529–533. 10.1016/j.psym.2016.05.001.

8. Malayala, S. V., Papudesi, B. N., Bobb, R., et al. Xylazine-Induced Skin Ulcers in a Person Who Injects Drugs in Philadelphia, Pennsylvania, USA. Cureus. 2022;14(8), e28160. 10.7759/cureus.28160.

9. Sisco E, Appley M. Identification of the veterinary sedative medetomidine in combination with opioids and xylazine in Maryland. J Forensic Sci. 2023;68(5):1708–1712. 10.1111/1556-4029.15242.

10. Dowton, A., Doernberg, M., Heiman, E., et al. Recognition and Treatment of Wounds in Persons Using Xylazine: A Case Report From New Haven, Connecticut. J Addict Med. 2023;17(6), 739–741. 10.1097/ADM.0000000000001198.

11. Zagorski CM, Hosey RA, Moraff C, et al. Reducing the harms of xylazine: clinical approaches, research deficits, and public health context. Harm Reduct J. 2023;20(1):141. 10.1186/s12954-023-00879-7.

12. D’Orazio, J., Nelson, L., Perrone, J., et al. Xylazine Adulteration of the Heroin-Fentanyl Drug Supply : A Narrative Review. Ann Intern Med. 2023;176(10), 1370–1376. 10.7326/M23-2001.

13. Mavragani, A., Ochoa, G., Tsagarakis, K. P. Assessing the Methods, Tools, and Statistical Approaches in Google Trends Research: Systematic Review. J Med Internet Res. 2018;20(11), e270. 10.2196/jmir.9366.

14. Rovetta A, Bhagavathula AS. Global infodemiology of COVID-19: analysis of Google web searches and Instagram hashtags. J Med Internet Res. 2020;25;22(8):e20673. 10.2196/20673.

15. Degenhardt, L., Bucello, C., Calabria, B., et al. What data are available on the extent of illicit drug use and dependence globally? Results of four systematic reviews. Drug Alcohol Depend. 2011;117(2-3), 85–101. 10.1016/j.drugalcdep.2010.11.032.

16. Young SD, Zheng K, Chu LF, et al. Internet searches for opioids predict future emergency department heroin admissions. Drug Alcohol Depend. 2018;190:166–169. 10.1016/j.drugalcdep.2018.05.009.

17. Patton, T., Abramovitz, D., Johnson, D., et al. Characterizing Help-Seeking Searches for Substance Use Treatment From Google Trends and Assessing Their Use for Infoveillance: Longitudinal Descriptive and Validation Statistical Analysis. J Med Internet Res. 2022;24(12), e41527. 10.2196/41527.

18. DEA. DEA Reports Widespread Threat of Fentanyl Mixed with Xylazine. Available from: https://www.dea.gov/alert/dea-reports-widespread-threat-fentanyl-mixed-xylazine. [Last Accessed on December 1, 2023].

19. Neumann, K., Mason, S. M., Farkas, K., et al. Harnessing Google Health Trends Data for Epidemiologic Research. Am J Epidemiol. 2023;192(3), 430–437. 10.1093/aje/kwac171.

20. Raubenheimer J. E. A practical algorithm for extracting multiple data samples from google trends extended for health. Am J Epidemiol. 2022;191(9), 1666–1669. 10.1093/aje/kwac088.

21. Altmetric. How is the Altmetric Attention Score calculated?. Available from: https://help.altmetric.com/support/solutions/articles/6000060969-how-is-the-altmetric-attention-score-calculated. [Last Accessed December 1, 2023].

22. Altmetric. When did Altmetric start tracking attention to each attention source?. Available from: https://help.altmetric.com/support/solutions/articles/6000136884-when-did-altmetric-start-tracking-attention-to-each-attention-source. [Last Accessed December 1, 2023].

23. FDA. FDA adverse event reporting system. Available from: https://open.fda.gov/data/faers/.html. [Last Accessed November 25, 2023].

24. Hafen R. geofacet: ‘ggplot2’ Faceting Utilities for Geographical Data. 2023. R package version 0.2.1. Available from: https://CRAN.R-project.org/package=geofacet. [Last Accessed October 10, 2023].

25. National Cancer Institute. Joinpoint regression program. Available from: https://surveillance.cancer.gov/joinpoint.htm. [Last Accessed August 21, 2023].

26. Irimata, K. E., Bastian, B. A., Clarke, T. C., et al. Guidance for Selecting Model Options in the National Cancer Institute Joinpoint Regression Software. Vital Health Stat 1, Programs and collection procedures. 2022;(194), 1–22.

27. Spadaro A, O’Connor K, Lakamana S, et al. Self-reported Xylazine Experiences: A Mixed-methods Study of Reddit Subscribers. J Addict Med. 2023;17(6):691–694. 10.1097/ADM.0000000000001216.

28. Palamar, J. J., Goldberger, B. A. Surveillance of Xylazine Use and Poisonings Is Needed-Without Blind Spots. Ann Intern Med. 2023;176(10), 1426–1427. 10.7326/M23-2299.

